# Relationship between social support and resilience among nurses: a systematic review

**DOI:** 10.1101/2022.09.04.22279592

**Authors:** Petros Galanis, Aglaia Katsiroumpa, Irene Vraka, Olga Siskou, Olympia Konstantakopoulou, Theodoros Katsoulas, Daphne Kaitelidou

## Abstract

**Background:** High levels of stress and anxiety in nurses negatively affect their physical and mental health and lead to poor job performance, limited job satisfaction, high levels of burnout and increased intention to leave the profession. By promoting resilience in nurses it is possible to reduce negative consequences for those working in highly stressful workplaces.

**Objective:** To summarize the evidence about the relationship between social support and resilience among nurses.

**Methods:** We performed a systematic review according to the Preferred Reporting Items for Systematic Reviews and Meta-Analysis (PRISMA) guidelines. We searched Pubmed, Medline, Scopus, Web of Science, and Cinahl from inception to August 28, 2022 using the following search strategy in all fields: ((resilience) AND (“social support”)) AND (nurses).

**Results:** Applying the inclusion criteria, we found six studies that investigated the relationship between social support and resilience among nurses. Two studies were conducted in China, two studies in Turkey, one study in Haiti, and one study in Taiwan. All studies were cross-sectionals and used convenience samples. All studies found a positive relationship between social support and resilience among nurses. In studies that used multivariable analysis, coefficient beta ranged from 0.13 to 0.69, while in studies that used correlation analysis, correlation coefficient ranged from 0.18 to 0.37.

**Conclusions:** We found that social support improves resilience among nurses. It is necessary to make systematic efforts to support nurses especially in the workplace. This need is even greater for inexperienced nurses, as it is more difficult for them to cope effectively with the difficulties of the profession. A harmonious working environment is essential to reduce the psychological pressure of nurses and improve job performance.

## Introduction

The ever-increasing professional demands on nurses, combined with the shortages in nursing staff in most countries, are leading nurses to considerable strain and often even burnout (Watson et al., 2008). This situation affects negatively both nurses and clients of healthcare services.

High levels of stress and anxiety in nurses negatively affect their physical and mental health and lead to poor job performance, limited job satisfaction, high levels of burnout and increased intention to leave the profession (Edwards et al., 2010; Rogers et al., 2004). Resilience can reduce stress, anxiety, burnout and intention to leave the profession, as well as has a positive effect on nurses’ physical and mental health (Cameron & Brownie, 2010; Matos et al., 2010; Mealer et al., 2012).

Resilience is defined as the ability of individuals to successfully cope with various stressful situations, particularly their ability to recover after highly traumatic situations such as the loss of a loved one (Jackson et al., 2007).

Resilience in the case of nurses is essential as it enables them to perceive meaning in their work and experiences and to mitigate the effects of the traumatic situations they face in the workplace (McAllister & McKinnon, 2009).

By promoting resilience in nurses it is possible to reduce negative consequences for those working in highly stressful workplaces (McCann et al., 2013). In fact, increased resilience plays an even more important role for nurses in the early years of their careers by significantly reducing their intention to leave the profession (Phillips et al., 2014).

Therefore, we conducted a systematic review to summarize the evidence about the relationship between social support and resilience among nurses.

## Methods

We searched Pubmed, Medline, Scopus, Web of Science, and Cinahl from inception to August 28, 2022 using the following search strategy in all fields: ((resilience) AND (“social support”)) AND (nurses). We performed the systematic review according to the Preferred Reporting Items for Systematic Reviews and Meta-Analysis (PRISMA) guidelines (Moher et al., 2009). We used the following inclusion criteria in our systematic review: (a) quantitative studies, (b) studies that report the relationship between social support and resilience, (c) studies that include nurses, (d) studies published in English. We collected the following data for each study included in the review: first author and year of publication, country, data collection time, sample size, age, study design, sampling method, and the main finding about the relationship between social support and resilience. The Joanna Briggs Institute critical appraisal tool was used to assess the risk of bias among the studies included in the review (Santos et al., 2018). According to this tool, the risk of bias was estimated as low, moderate and high.

## Results

The flow diagram of the systematic review according to PRISMA guidelines is shown in Figure 1. After the first search in the databases and the removal of the duplicate records we found 13,232 records. Applying the inclusion criteria, the total number of studies that were included in the review was six.

**Figure 1.**
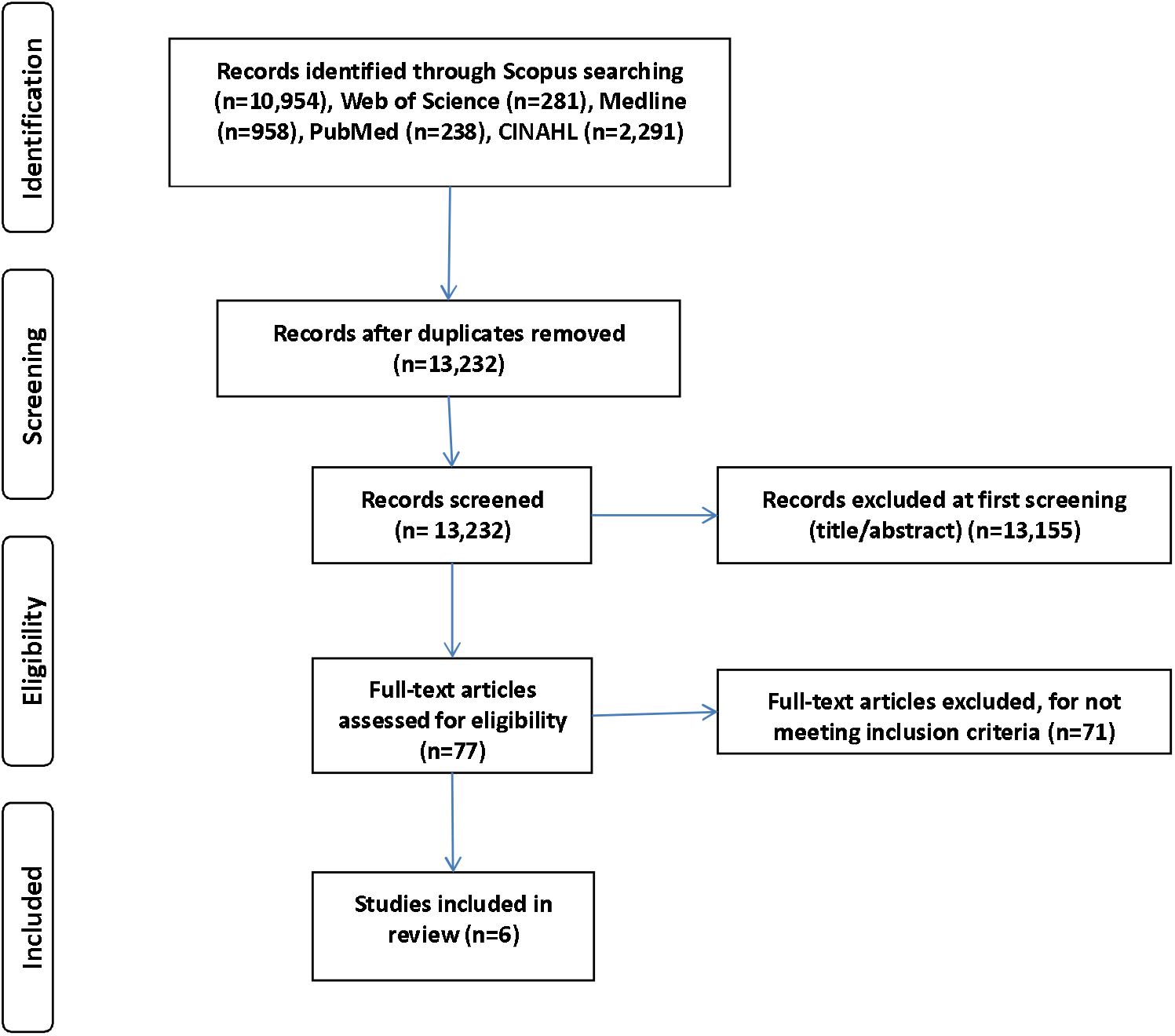
The flow diagram of the systematic review according to PRISMA guidelines.

Main characteristics of the six studies included in the systematic review are shown in Table 1. Two studies were conducted in China (Liu et al., 2018; Wang et al., 2018), two studies in Turkey (Kılınç & Sis Çelik, 2021; Öksüz et al., 2019), one study in Haiti (Caton, 2021), and one study in Taiwan (Hsieh et al., 2017). Studies were conducted through 2013-2020. The minimum sample size was 50 nurses, while the maximum sample size was 747 nurses. All studies were cross-sectionals and used convenience samples. Risk of bias of studies is shown in Table 2. All studies had a low level of bias.

**Table 1.** Main characteristics of the studies included in the systematic review.

| First author and year of publication | Country | Data collection time | Sample size (n) | Age, mean (standard deviation) | Study design | Sampling method | Main findings |
| --- | --- | --- | --- | --- | --- | --- | --- |
| (Caton, 2021) | Haiti | September 2018 to January 2019 | 50 | 43 (5.7) | Cross-sectional | Convenience sample | Multivariable analysis showed that social support increases resilience (coefficient beta = 0.69, p-value < 0.001) |
| (Hsieh et al., 2017) | Taiwan | June to December 2013 | 230 | ≤30 years, 61.7%; >30 years, 38.3% | Cross-sectional | Convenience sample | Multivariable analysis showed that family support (coefficient beta = 0.13, p-value = 0.031) and peer support (coefficient beta = 0.17, p-value = 0.006) increase resilience |
| (Kılınç & Sis Çelik, 2021) | Turkey | April 2020 | 370 | ≤40 years, 91.1%; >40 years, 8.9% | Cross-sectional | Convenience sample | Correlation analysis showed that family support (r = 0.34, p-value < 0.001), friends support (r = 0.37, p-value < 0.001), and support by significant others (r = 0.38, p-value < 0.001) was related positively with resilience |
| (Liu et al., 2018) | China | May 2017 | 183 | 34.1 (7.2) | Cross-sectional | Convenience sample | Correlation analysis showed that social support was related positively with resilience (r = 0.22, p-value < 0.01) |
| (Öksüz et al., 2019) | Turkey | November 2015 to February 2016 | 242 | ≤30 years, 42.2%; >30 years, 57.8% | Cross-sectional | Convenience sample | Correlation analysis showed that social support was related positively with resilience (r = 0.18, p-value = 0.006) |
| (Wang et al., 2018) | China | August to November 2015 | 747 | 23.5 (1.7) | Cross-sectional | Convenience sample | Multivariable analysis showed that coworker support (coefficient beta = 0.13, p-value = 0.017) increases resilience |

**Table 2.** Quality of studies included in this systematic review.

|  | (Caton,<br>2021) | (Hsieh et<br>al., 2017) | (Kılınç & Sis<br>Çelik, 2021) | (Liu et<br>al., 2018) | (Öksüz et<br>al., 2019) | (Wang et<br>al., 2018) |
| --- | --- | --- | --- | --- | --- | --- |
| 1. Were the criteria for inclusion in the sample clearly defined? | Yes | Yes | No | No | No | Yes |
| 2. Were the study subjects and the setting described in detail? | Yes | Yes | Yes | Yes | Yes | Yes |
| 3. Was the exposure measured in a valid and reliable way? | Yes | NA | Yes | Yes | Yes | NA |
| 4. Were objective, standard criteria used for measurement of the condition? | Yes | Yes | Yes | Yes | Yes | Yes |
| 5. Were confounding factors identified? | Yes | NA | NA | NA | NA | NA |
| 6. Were strategies to deal with confounding factors stated? | Yes | NA | NA | NA | NA | NA |
| 7. Were the outcomes measured in a valid and reliable way? | Yes | Yes | Yes | Yes | Yes | Yes |
| 8. Was appropriate statistical analysis used? | Yes | Yes | Yes | Yes | Yes | Yes |
| **Risk of bias** | **Low** | **Low** | **Low** | **Low** | **Low** | **Low** |

NA: not applicable

All studies found a positive relationship between social support and resilience among nurses. Three studies measured social support in general (Caton, 2021; Liu et al., 2018; Öksüz et al., 2019), while the other three studies measured family support, friends support, and coworkers support (Hsieh et al., 2017; Kılınç & Sis Çelik, 2021; Wang et al., 2018). Three studies used multivariable analysis to eliminate confounders (Caton, 2021; Hsieh et al., 2017; Wang et al., 2018), while the other three studies found the correlation coefficient between social support and resilience without eliminating confounding (Kılınç & Sis Çelik, 2021; Liu et al., 2018; Öksüz et al., 2019). In studies that used multivariable analysis, coefficient beta ranged from 0.13 to 0.69, while in studies that used correlation analysis, correlation coefficient ranged from 0.18 to 0.37.

## Discussion

We conducted a systematic review to found the relationship between social support and resilience among nurses. We identified six studies that were conducted in four countries; China, Turkey, Haiti and Taiwan. All studies found a positive relationship between social support and resilience.

The support that nurses receive proves to be a key parameter in enhancing their resilience, as it enables nurses to manage more effectively the negative emotions they experience due to their work (Kılınç & Sis Çelik, 2021). More specifically, nurses work under highly stressful conditions and support from others reinforces their belief that they are not alone in dealing with their problems, but instead can find support and help.

In fact, the more support nurses receive, the better the way they manage the stress they experience. Moreover, greater support enables nurses to better adapt to demanding circumstances both at the professional and personal level. In this case, it is necessary to have an in-depth understanding of the nurses’ family, friends and professional environment so that we can provide them with the best possible support network. This will enable nurses to strengthen their resilience, face their work with more enthusiasm and better manage the various mental health issues (Liu et al., 2018).

Support from family is particularly important, as it reduces the feeling of loneliness in dealing with problems and makes it easier to manage problems (Fu et al., 2018; Orgambídez-Ramos & de Almeida, 2017). In addition, support from family or relatives has a positive impact on the physical and mental health of nurses working in hospitals (Fu et al., 2018; Sun et al., 2017).

Moreover, support from coworkers increase resilience through an indirect effect of self-efficacy (Wang et al., 2018). This kind of support is very important for early career nurses since they must adapt in a difficult work environment. Supportive work environment strength employees and improve job performance. Colleagues can significantly help inexperienced nurses to cope more effectively with difficult clinical situations as well as professional challenges by providing them with emotional support, advice and professional help in the hospital setting (Welsh, 2014). Peer support can alleviate stress, reduce psychological problems and increase the likelihood of young nurses staying in the profession (Hall, 2007).

It is noted that some categories of nurses have reduced resilience. For example, nurses working in psychiatric hospitals, nurses who do not have a permanent employment relationship with the hospital where they work and nurses who work more often on a rotational basis have more problems due to their reduced resilience (Liu et al., 2018). It is also important that women have lower resilience than men (Aburn et al., 2016; Öksüz et al., 2019). Therefore, special attention needs to be paid to these categories of nurses, especially by motivating nurses and strengthening their ambitions for a better professional future.

Our systematic review had several limitations. First, we searched in five databases using specific keywords. Second, all studies were cross-sectional and thus a causal relationship between social support and resilience could not be established. Third, three out of six studies did not multivariable analysis to eliminate confounding. Forth, the number of studies was low. Fifth, three studies calculated a coefficient beta and three studies calculated a correlation coefficient. Thus, we cannot perform a meta-analysis to provide a pooled estimation of the relationship between social support and resilience due to the low number of studies.

In conclusion, social support improves resilience of nurses. Therefore, it is necessary to make systematic efforts to support nurses especially in the workplace. This need is even greater for inexperienced nurses, as it is more difficult for them to cope effectively with the difficulties of the profession. For instance, nurse managers in hospitals could arrange workshops, talk of the trouble, training sessions, working group activities, regular meetings, and seminars to improve resilience among nurses. A harmonious working environment is essential to reduce the psychological pressure of nurses and improve job performance.

## Data Availability

All data produced in the present study are available upon reasonable request to the authors

## Notes

### Competing Interest Statement

The authors have declared no competing interest.

### Funding Statement

This study did not receive any funding

